# Is it possible to flatten-the-curve after the initial outbreak of Covid-19? A data-driven modeling analysis for Omicron pandemic in China

**DOI:** 10.1101/2022.12.21.22283786

**Authors:** Jiaqi Sun, Yusi Li, Ning-Yi Shao, Miao Liu

## Abstract

In the current coronavirus disease 2019 (COVID-19) pandemic, the Omicron variant of severe acute respiratory syndrome coronavirus 2 has become the predominant strain circulating worldwide. In China, enormous controversies exist regarding the “dynamic zero tolerance” (DZT) and “totally no inventions” (TNI) strategies for preventing the spread of the Omicron variant. Currently, China is gradually relaxing the COVID-19 measures from DZT level. In such situations, the “flatten-the-curve” (FTC) strategy, which decreases and maintains the low rate of infection to avoid overwhelming the healthcare system by adopting relaxed nonpharmaceutical interventions (NPIs) after the initial outbreak, has been perceived as most appropriate and effective method to prevent the spread of the Omicron variant. Hence, we established a data-driven model of Omicron transmission based on the pandemic data of Macau, Hong Kong, and Singapore in 2022 to deduce the overall prevention effect throughout China. In the current immunity level without any NPI applied, more than 12.7 billion (including asymptomatic individuals) were infected with the Omicron variant within 90 days, but the daily new infections sharply declined; moreover, Omicron outbreak would result to 1.49 million deaths within 180 days. The application of FTC could decrease the deaths by 36.91% within 360 days. Age-stratified analyses showed that the NPI application among individuals aged >60 years would also result in 0.81 million deaths within 360 days, and the application of FTC strategy through treatment with anti-COVID drugs can reduce the number of deaths to 0.40 million. In a model of completed vaccination, the application of TNI strategy would also result in 0.56 million deaths and slightly decrease the infection numbers. The strict implementation of FTC policy combined with completed vaccination and drug use, which only resulted in 0.19 million deaths in an age-stratified model, will help end the pandemic within about 240 days. The pandemic would be terminated within a shorter period of time without resulting in a high fatality rate; therefore, the FTC policy could be strictly implemented through enhancement of immunity and drug use.

## Introduction

Since the COVID-19 outbreak occurred in late December 2019, the severe acute respiratory syndrome coronavirus 2 (SARS-Cov-2) has constantly mutated into multiple variants that have been identified to date^1^. The Omicron variant has been reported to have several subtypes since its discovery in 2021 and has been the dominant strain circulating worldwide^2^. Even in China, despite the strict implementation of massive anti-epidemic measures, Omicron flare-ups occasionally occurred; therefore, the nationwide surge of Omicron is still imminent^3^. Although current data suggests that Omicron has a surprisingly high rate of transmission, rapid replication in the human bronchus, and strong immune escape potential, it does not seem to cause more severe disease compared with other SARS-Cov-2 variants^4^. The high rate of asymptomatic infection, mild impairment of the lungs and other organs, and short recovery time brought into doubt the necessity of epidemic prevention and control with strict implementation of nonpharmaceutical interventions (NPIs)^5-7^.

The low infection rate in China was attributed to the continuous adoption of the dynamic zero tolerance policy^8^. In the ongoing Omicron-dominant pandemic, the specific policy that should be applied remains controversial. In some countries, the negative impact of COVID-19 has somehow reduced; however, although less infection rates were achieved, the zero-COVID policy is still applied in China^9^. Completely eradicating the pandemic within the shortest possible time is the safest method to protect the older population as the application of high-risk policy will further prolong the pandemic. According to a previous mathematical model research, although >90% of the population in China had already been vaccinated, more than 1.5 million people will die of COVID-19 if all NPIs are discontinued while still confronting Omicron transmission; considering the Chinese population’s current immune level, majority of them is expected to develop the infection within 6 months^10^. The results of this model are difficult to apply in a real-life setting. Therefore, a less stringent, non-lockdown, but effective strategy is required to curb the transmission of Omicron; however, the stringency between DZT and TNI is considered to compromise the solution. Typically, the number of infected persons in the COVID-19 outbreak suddenly increases during the initial period, which is illustrated as a steep peak-like curve in a statistical graph. The implementation of mild NPI measures can decrease the current infection rates; hence, the curve could be flattened for a longer time, and the number of cases will gradually decline, resulting in a fewer number of new infection cases. This degree of prevention was referred to as “flatten-the-curve” response to pandemic, which permits virus transmission at a low level without affecting the overall healthcare system^11,12^. Unduly harsh policy may cause inconvenience and huge financial burden. China has the highest proportion of older adults among the countries worldwide, and a large part of this population with underlying health conditions are more vulnerable to COVID-19^13-16^. Therefore, total liberalization of intervention programs to control the pandemic is suspected to increase the risk for severe disease or death from Omicron virus. Compared with the “all-or-none” response strategy, FTC policy is aimed at maintaining the social homeostasis, avoiding the possible collapse of the healthcare system, and reducing mortality^11,12^.

To date, no study has evaluated the exact effect of FTC. Whether the FTC policy is the most effective strategy to cope with the Omicron epidemic and the kinds of drugs and the population protective immunity level needed to stop the pandemic remain unclear. In this study, we established a data-driven model, based on the progress of Omicron outbreak in Macau, Hong Kong, and Singapore in 2022, to evaluate the possibility of adopting the FTC policy in future pandemics in China.

## Methods

SARS-CoV-2 Omicron transmission, vaccination rate, and disease burden The baseline model used in our study was improved based on the age-structured stochastics compartmental susceptible-latent-infectious-removed-susceptible model developed by the research team from Shanghai, China^10^. In the baseline scenario, 5 imported infections were used in the baseline simulations performed for 12 months. Then, 20 imported infections were included in the sensitivity analysis (Figure S1). The basic reproduction number (R) of the Omicron subtype was estimated to more than 3-fold times of Delta variant (from 3 to 8)^17, 18^. The improvements in our model were as follows: 1. the effect of herd immunity was considered in our model, as the immunity was (*x*+1)^2^, as *x* is the infected population; 2. the scenario about reducing effective contacts with different intensity levels among different age groups was analyzed.

Then, the number of infections, confirmed cases, hospitalizations, intensive care unit (ICU) admissions, and deaths were calculated to assess the SARS-CoV-2 Omicron burden. The parameters used in the mathematical model for inferring the above index on SARS-CoV-2 Omicron progression were the same as that used by the Shanghai research team^10^. With regard to the healthcare resources, a total of 9.1 million hospital beds were utilized; of these, 3.14 million beds were allocated for patients with respiratory illness admitted in the internal medicine department, pediatric department, infectious disease department, and ICUs in China, of which 64,000 belonged to ICU^19^.

### Mitigation with vaccination, antiviral therapies, and NPIs

Inactivated vaccines were administered, and the proportion of vaccination in the different age groups were the same as that reported by the Shanghai research team^10^. The vaccine efficacy on infection, onward transmission rate, symptoms, hospitalization rate, vaccination transition rate, and recovery rate were the same as that reported in a previous study^10^. We adjusted the vaccine effectiveness against COVID-19-related death, as shown in Table S1.

In the baseline scenario, no antiviral therapies were provided to the patients with confirmed cases. To quantify the mitigation effect of antiviral therapies, we simulated another scenario; approximately, 75% of patients with confirmed cases were treated with PAXLOVID in accordance with the Diagnosis and Treatment Protocol for Novel Coronavirus Pneumonia (Trial Version 7) issued by the Chinese Center for Disease Control and Prevention^20^. The effectiveness rate of the antiviral therapies was assumed to be 75%.

The intensity of NPI was rated from 0 to 1. The NPI intensity level (NPIIL) ranged from 0 to 1. An NPIIL level 0.1 indicated that the strictest NPI was implemented, while level 1 indicated that no NPI was implemented. In the baseline scenario, no NPI was carried. To quantify the mitigation effect of NPIs, we simulated two alternative scenarios: 1. reducing the effective contacts with the same intensity level among different age groups and 2. reducing the effective contacts with different intensity levels among different age groups.

To evaluate the effect of the positive scenario, we used the Singapore scenario as a reference; the completed mRNA vaccine efficacy is shown in Table S1. The definition of completed vaccine was adopted from the standards of Ministry of Health in Singapore^21^. In the positive scenario, the proportion of older adults aged ≥60 years who could not receive vaccination was only 0.1. Hence, we assumed that 75% of patients with confirmed cases received PAXLOVID. The SARS-CoV-2 Omicron burden was predicted at specific times among different age groups using different NPIIL of NPI mitigation strategies based on the positive scenario (Figure S3).

### Statistical analysis

A total of 200 stochastic simulations were performed for each scenario. The results of these simulations determined the distribution of infections, confirmed cases, hospitalizations, ICU admissions, and deaths by age. Then the median and 2.5% and 97.5% quantiles of 200 simulations were calculated.

### Data availability and code availability

All data used in this modeling study were collected from publicly available databases. The data used in the study are provided in the supplementary materials, and the codes are available on GitHub.

## Results

### Baseline scenario

In the baseline scenario, NPI protocols for the prevention of SARS-CoV-2 Omicron transmission were not applied, and antiviral therapies for controlling the progress of disease were not administered. The simulated conditions in our model were based on the age-structured stochastic compartmental susceptible-latent-infectious-removed-susceptible model of SARS-CoV-2 transmission. The different values presented between our model and the previous model indicates the transmission rate; moreover, herd immunity was introduced in our model: 1) the transmission rate in the absence of NPIs, which was inferred from the reproduction number of the SARS-CoV-2 Omicron variant, was set at 7.2, and 2) herd immunity was determined based on the proportion of daily infectious cases.

The predicted daily SARS-CoV-2 Omicron burden in China according to the baseline scenario is shown in Figure 1. The total number of infectious cases and accumulated number of deaths in the total population were 1273.27 million (close to 12.7 billion when came to 90 days) and 1.49 million, respectively. The total number of infectious cases and accumulated number of deaths among the older group were 244.60 million and 1.40 million, respectively, accounting for 19.21% of the total number of infectious cases and 93.97% of the total accumulated deaths. On day 52, the total daily infectious cases reached 92.29 million, among which 17.56 million (19.03%) were older adults. The total daily deaths peaked on day 59 (72,093), and 68,122 (94.45%) of whom were older adults. On day 58, the numbers of non-ICU patients and ICU patients surged to 1.67 million and 1.07 million, respectively. Moreover, 1.05 million and 0.98 million older adults required to stay in the hospital and ICU, respectively, accounting for 62.64% and 91.57% of the total number of non-ICU and ICU patients, respectively. From day 44 to day 88, the required ICU beds exceeded the number of daily ICU beds available in China.

**Figure 1.**
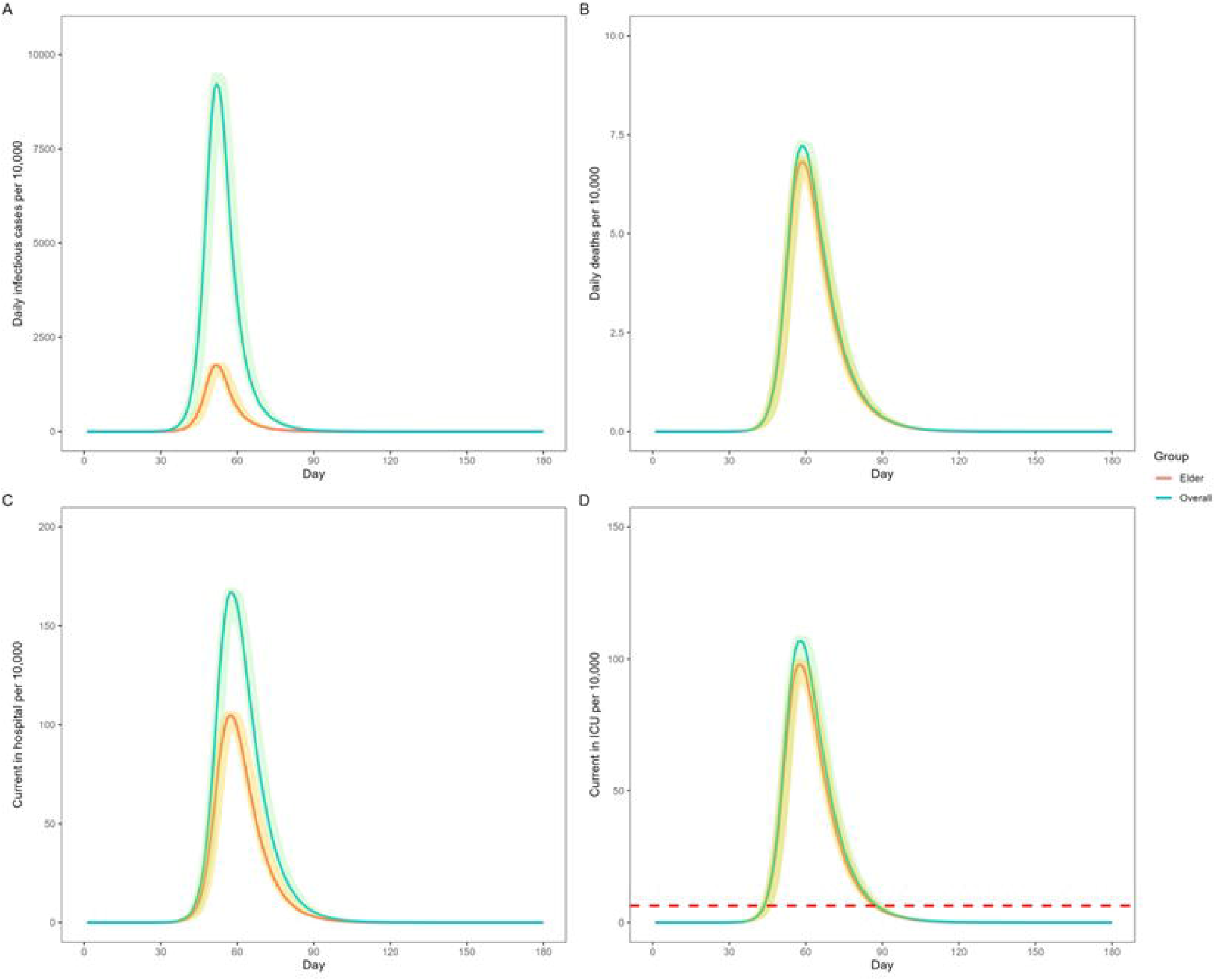
Predicted SARS-CoV-2 Omicron burden in China with the baseline scenario. Note: A. Daily number of infectious cases per 10,000 individuals B. Daily number of deaths per 10,000 individuals C. Current number of non-ICU patients per 10,000 individuals D. Current number of ICU patients per 10,000 individuals In D, the red dashed line indicates the number of ICU beds available in China. All data are presented as median with 2.5% and 97.5% quantiles of n=200 simulations. ICU, intensive care unit

### Classification of the stringency of NPIs

The NPI stringency was rated on a scale of 0–1. A NPI stringency level (NPISI) 0.1 indicated that the strictest NPI was carried out, while NPISI 1 indicated that no NPI was implemented. Our baseline scenario model is used to predict daily the number of infectious cases caused by SARS-CoV-2 Omicron variant in Macau. Here, we supposed that 5 initial infectious were introduced. The daily predictive and actual SARS-CoV-2 Omicron burden in Macau are shown in Figure S2. After comparing the real-world data with the predicted data, we adjusted the NPI level parameters in the model to determine the specific NPIs of the NPISI in the model that corresponds to the real-world situation. The application of FTC could decrease the deaths by 36.91% within 360 days. (Table 1 and Figure S2).

**Table 1.** Classification of the NPIIL

| NPIIL | NPIs |
| --- | --- |
| 0.80 | Wearing of facemask |
| 0.30 | Banning from dining in a restaurant and closure of entertainment venues and other public places |
| 0.05 | Restriction on going outdoors |
| 0.02 | Requirement for rapid antigen testing on a daily basis and quarantining of patients with confirmed cases |

NPIs: nonpharmaceutical interventions
NPIIL: nonpharmaceutical intervention intensity level
NPIILs between 0 and 1. NPIIL level 0.1 indicates that the strictest NPI was carried out, while NPIIL level 1 indicated that no NPI was implemented.

### Impact of NPI mitigation strategies

We carried out different NPISI of NPI mitigation strategies at specific timepoints to avoid the shortage of ICU beds. The following two subset scenarios were separately analyzed: (1) all patients with confirmed cases did not receive drug treatment, and (2) 75% of patients with confirmed cases received drug treatment. The efficacy of the drug was assumed to be 75%.

Results of the prediction of daily ICU beds required using different NPIIL of NPI mitigation strategies at specific timepoints are shown in Figure 2. In all patients with confirmed cases who did not receive PAXLOVID, the NPIs were carried out on day 1, day 40, day 170, and day 270 with the NPIILs of 0.8, 0.2, 0.3, and 0.5, respectively (Figure 2A). The number of daily ICU beds required peaked on day 127 (65), and a total of 197 ICU beds were needed (Figure 2A). For older patients, the daily ICU beds needed on day 127 was 60,923 (93.46%) (Figure 2A). The total number of infections in 360 days was 680.40 million, of whom 134.45 million (19.76%) were older patients (Figure 2B). The total number of deaths in 360 days was 0.94 million, of whom 0.89 million (95.25%) were older patients (Figure 2C).

**Figure 2.**
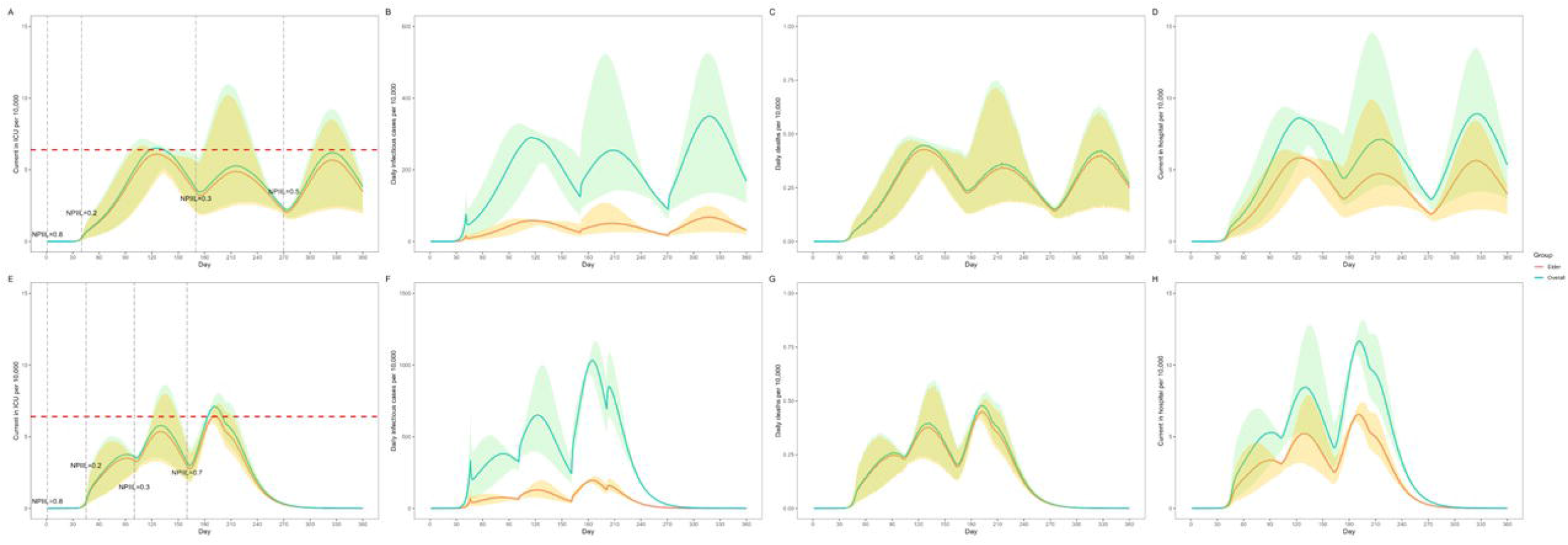
Prediction of SARS-CoV-2 Omicron burden using different NPIILs of NPI mitigation strategies at specific timepoints. Note: A. Current number of ICU patients per 10,000 individuals who did not receive PAXLOVID B. Daily number of infectious cases per 10,000 individuals who did not receive PAXLOVID C. Daily number of deaths per 10,000 individuals who did not receive PAXLOVID D. Current number of hospitalized (non-ICU) patients per 10,000 individuals who did not receive PAXLOVID E. Current number of ICU patients per 10,000 individuals with 75% of patients with confirmed cases receiving PAXLOVID F. Daily number of infectious cases per 10,000 individuals with 75% of patients with confirmed cases receiving PAXLOVID. G. Daily number of deaths per 10,000 individuals with 75% of patients with confirmed receiving PAXLOVID. H. Current number of hospitalized (non-ICU) patients per 10,000 individuals with 75% of patients with confirmed cases receiving PAXLOVID In A and E, the red dashed line indicates the number of ICU beds available in China. NPIIL: nonpharmaceutical intervention intensity level NPIILs between 0 and 1. NPIII level 0.1 indicates that the strictest NPI was carried out, while NPIII level 1 indicated that no NPI was implemented. All data are presented as median with 2.5% and 97.5% quantiles of n=200 simulations.

In 75% of the patients with confirmed cases who received PAXLOVID, the NPIs were carried out on days 1, 45, 100, and 160 with NPIILs of 0.8, 0.2, 0.3, and 0.7, respectively (Figure 2E). The daily ICU bed needed peaked on day 192 (71,037 ICU beds) (Figure 2E). For older patients, the daily ICU beds needed on day 192 were 64,577 (90.91%) (Figure 2E). The total number of infections in 360 days was 1053.20 million, of whom 204.25 million were older patients (19.39%) (Figure 2F). The total number of deaths in 360 days was 0.58 million, of whom 0.55 million (94.44%) were older patients (Figure 2G).

### Impact of NPI mitigation strategies by age

We investigated the predicted daily SARS-CoV-2 Omicron burden with different NPIILs of NPI mitigation strategies at specific timepoints among different age groups under the following two conditions: (1) all the confirmed cases did not receive PAXLOVID, and (2) 75% of the patients with confirmed cases received PAXLOVID.

Different NPIILs of NPI mitigation strategies at specific timepoints among different age groups were set to avoid the shortage of ICU beds. The NPIIL was separately defined in three contact groups: “inside <60 group (contact between the population aged <60 years),” “between group (contact between the population aged <60 years and the older adults aged ≥60 years),” and “inside ≥60 group (contact between older adults aged ≥60).”

In all patients with confirmed cases who did not receive PAXLOVID, the NPIs were conducted on days 1, 14, 45, 150, 220, and 270 with different NPIILs in the “inside < 60 group,” “between group,” and “inside ≥ 60 group” (Figure 3A). The number of daily ICU beds needed peaked on day 200 (66,020 ICU beds) (Figure 3A). For older patients, the daily ICU beds needed on day 200 were 54,728 (82.90%) Figure 3A). The total number of infections in 360 days was 990.67 million, of whom 103.59 million (10.46%) were older adults (Figure 3B). The total number of deaths in 360 days was 0.81 million, of whom 0.74 million (90.88%) were older adults (Figure 3C).

**Figure 3.**
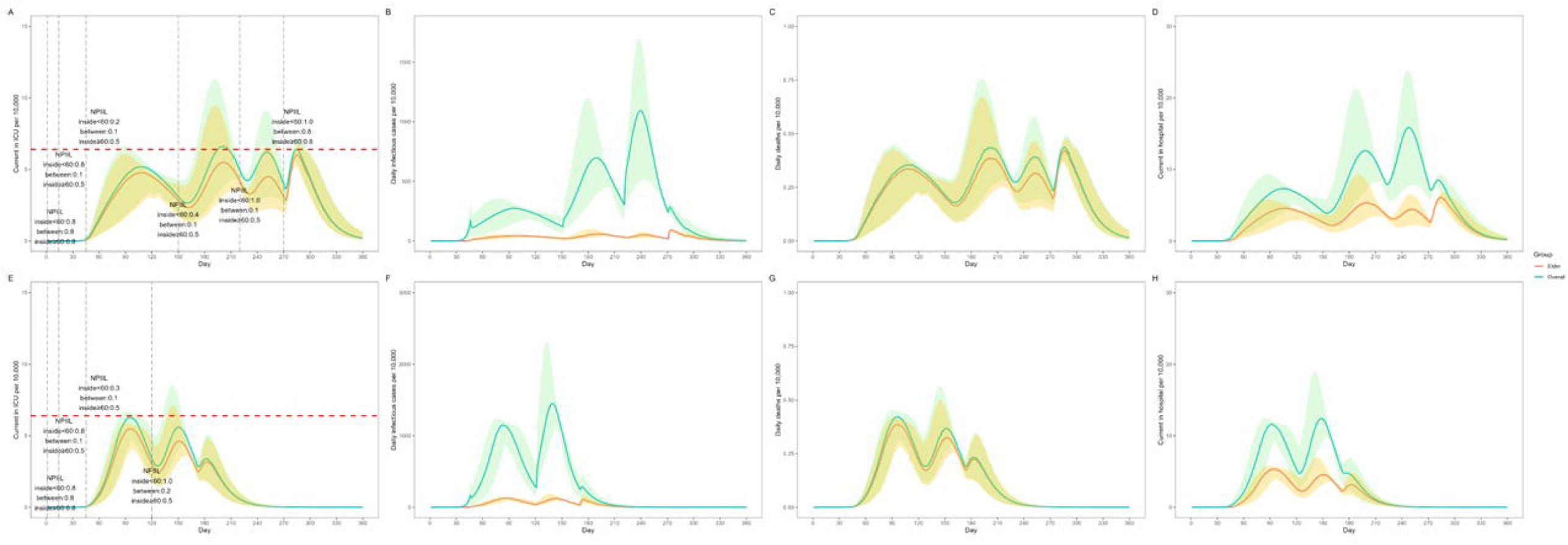
Prediction of SARS-CoV-2 Omicron burden using different NPIILs of NPI mitigation strategies at specific timepoints among different age groups Note: A. Current number of ICU patients per 10,000 individuals who did not receive PAXLOVID B. Daily number of infectious cases per 10,000 individuals who did not receive PAXLOVID. C. Daily number of deaths per 10,000 individuals who did not receive PAXLOVID D. Current number of hospitalized (non-ICU) patients per 10,000 individuals who did not receive PAXLOVID. E. Current number of ICU patients per 10,000 individuals with 75% of patients with confirmed cases receiving PAXLOVID F. Daily number of infectious cases per 10,000 individuals with 75% of patients with confirmed cases receiving PAXLOVID G. Daily number of deaths per 10,000 individuals with 75% of patients with confirmed cases receiving PAXLOVID H. Current number of hospitalized (non-ICU) patients per 10,000 individuals with 75% of patients with confirmed cases receiving PAXLOVID In A and E, the red dashed line indicates the number of ICU beds available in China. NPIIL: nonpharmaceutical intervention intensity level NPIILs between 0 and 1. NPIII level 0.1 indicates that the strictest NPI was carried out, while NPIII level 1 indicated that no NPI was implemented. Inside <60 group: contact between the population aged < 60 years, between: contact between the population aged <60 years and older adults aged ≥60 years; inside ≥60: contact between older patients aged ≥60 years All data are presented as median with 2.5% and 97.5% quantiles of n=200 simulations.

In 75% of the patients with confirmed cases who received PAXLOVID, the NPIs were conducted on days 1, 14, 45, and 120 with different NPIILs in the “inside < 60 group,” “between group,” and “inside ≥ 60 group” (Figure 3E). The daily ICU beds peaked on day 94 (62,888 ICU beds) (Figure 3E). In older patients, the daily ICU beds needed on day 94 were 54,798 (87.14%) (Figure 3E). The total number of infections in 360 days was 1019.30 million, of whom 119.39 million (11.71%) were older patients (Figure 3F). The total number of deaths in 360 days was 0.40 million, of whom 0.37 million (91.47%) were older patients (Figure 3G).

### Comparison of the outcomes of different strategies

We also assumed an ideal scenario in which different age-level groupings of NPI was implemented, 75% of the patients took Paxlovid, and the mortality rate decreased to 0.025% after vaccination (based on data obtained from Singapore). We explored the accumulated number of infections and deaths in the total population and older group following the use of different strategies.

The accumulated infections and accumulated deaths of the total population and older adults in 360 days in different models are shown in Figure 4 and Table 2. In Table 2, the prevalence and fatality of SARS-CoV-2 Omicron in the total population and older adults were also calculated. In Model 5, in which 75% of patients with confirmed cases received PAXLOVID and different NPIILs of NPI mitigation strategies applied among different age groups at specific timepoints were set to avoid the shortage of ICU beds, the total number of infectious cases was 1.41 billion, accounted for 72.19% of the total number of people in the country; meanwhile, the total number of infectious older adults was 267.36 million, accounting for 44.66% of the total number of older people in the country. The fatality rates were 0.04% in the total population and 0.31% in the older group. When the restrictions were fully lifted after 180 days, 170,966 people died after 180 days, of whom 159,268 were older adults. In the ideal model, the prevalence rates of Omicron infection were 71.66% in the total population and 59.67% in the older adults. The fatality rates in the total population and older adults were 0.02% and 0.11%, respectively. When the restrictions were fully lifted 180 days later, 61,957 people died after 180 days, of whom 51,909 were older adults.

**Table 2.** Accumulated number of infectious cases and accumulated number of deaths caused by SARS-CoV-2 Omicron in different models.

| Model | Overall |  | Older patients |  |
| --- | --- | --- | --- | --- |
|  | Accumulated number of infectious cases | Accumulated number of deaths | Accumulated number of infectious cases | Accumulated number of deaths |
| Model 1: baseline scenario (TNI) | 1273.27<br>(90.16%) | 1.49 (0.12%) | 244.60<br>(91.49%) | 1.40 (0.57%) |
| Model 2: NPIs | 680.40<br>(48.19%) | 0.94 (0.14%) | 134.45<br>(50.29%) | 0.89 (0.66%) |
| Model 3: NPIs + 75% of cases with PAXLOVID | 1053.20<br>(74.59%) | 0.58 (0.06%) | 204.25<br>(76.40%) | 0.55 (0.27%) |
| Model 4: NPIs by age-stratified | 990.67<br>(70.16%) | 0.81 (0.08%) | 103.59<br>(38.75%) | 0.73 (0.70%) |
| Model 5: TNI + completed vaccination on 90% of population | 1259.43<br>(89.19%) | 0.56 (0.04%) | 237.69<br>(88.90%) | 0.51 (0.21%) |
| Model 6: NPIs by age-stratified + 75% of cases with PAXLOVID | 1019.30<br>(72.19%) | 0.40 (0.04%) | 119.39<br>(44.66%) | 0.37 (0.31%) |
| Model 7: completed vaccination on 90% of population + 75% of cases with PAXLOVID + NPIs by age-stratified | 1011.87<br>(71.66%) | 0.19 (0.02%) | 159.53<br>(59.67%) | 0.17 (0.11%) |

The total number of people is 1.41 billion and the total number of elders is 267.36 million in China.

**Figure 4.**
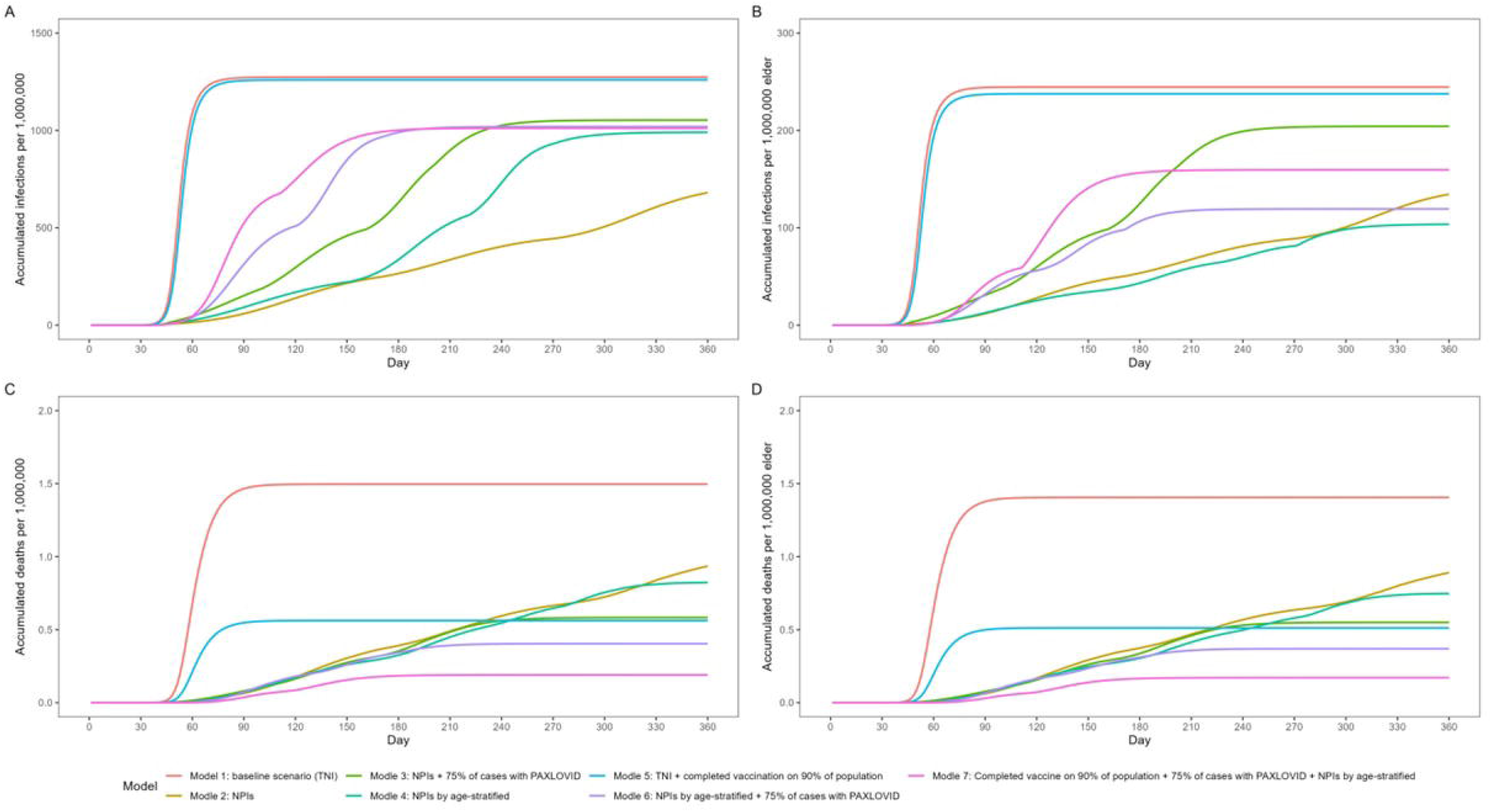
Comparison of the accumulated number of infectious cases and accumulated number of deaths caused by SARS-CoV-2 Omicron in different models. Note: A. The overall accumulated number of infectious cases of SARS-CoV-2 Omicron in different models B. The accumulated number of infectious cases caused by SARS-CoV-2 Omicron among older adults in different models C. The overall accumulated number of deaths caused by SARS-CoV-2 Omicron in different models D. The accumulated number of deaths caused by SARS-CoV-2 Omicron among older adults in different models

The greatest number of fatal falls occurs if the baseline status peaks within a sharp time. The least number of deaths was reported in the ideal scenario; hence, we recommend the following model: “PAXLOVID treatment in 75% of the population + application of age-stratified FTC + receipt of complete vaccine (3 doses of mRNA) in 90% of the population.” The number of deaths in this model was less than 200,000 when came to the peak value (within 240 days); hence, these findings show that the pandemic has a tendency to end within a shorter period of time. In other models, the number of deaths reported were several times greater.

## Discussion

Although the transmission model based on Shanghai’s data made some predictions of the Omicron transmission in China, some conclusions in this study were not applicable in the real-world situation. According to their inferences, the total numbers of infected people were significantly high compared with that of the symptomatic people (more than 2 billion), which is beyond the actual population size^10^. In the current study, we corrected and optimized the modelling system and followed the epidemiological data reports in Macau, Hong Kong, and Singapore in 2022. We also determined whether the FTC policy was an effective method for the prevention and control of the Omicron outbreak. Here, we modified the model to revise the effect of the herd immunity (see Methods), as the revised model indicated that the basal scenario will reach 1.27 billion accumulated patients at day 180.

Our results also proved the kind of FTC prevention applied, at least several hundred thousand of deaths may occur. According to the conclusion of other studies, the insufficient immunity level of the Chinese population was one of the reasons for the continued implementation of the DZT policy^10^.

COVID-19 mixed vaccines or bivalent COVID-19 vaccines contribute to the significant increase in the antibody levels and provide broad protection for most of the populations; therefore, multiple sources of vaccines are an important prerequisite to end the pandemic^22, 23^. The promotion of immunity through vaccination along with the implementation of FTC policy would effectively overcome the COVID-19 outbreak within a shorter period of time.

The implementation of FTC policy permits virus transmission in a relatively low level; however, the Omicron variant is one of the most highly infectious viruses; although the case fatality rate was close or lower than that of influenza, the high number of infected individuals would result in the increased number of deaths^10^. Promisingly, the Omicron variant has a low pathogenicity; therefore, patients with asymptomatic cases and mild symptoms do not require hospital admission, to avoid exhausting the healthcare system. In outbreaks caused by a virus with extremely high transmissibility, like Omicron, the implementation of non-DZT policy is not effective in preventing and controlling its spread among individuals of all ages. The national health code system for every citizen had been established in China, which provided data on age, sex, and movement of people in real time^24^. This system contributes to the implementation of age-stratified FTC.

However, this study has some limitations. China had increased the ICU bed capacity, but we only used 64,000 beds in this study. Owing to the difference in vaccination types, the actual numbers of deaths may have been underestimated. In our research, FTC should be combined with Paxlovid use and mRNA vaccination in order to reduce the number of deaths. Paxlovid is an oral drug for patients with mild COVID-19 to avoid progression to severe COVID-19, which requires hospitalization^25^. The scope of application (75%) in our model is reasonable. Although 90% of the population received inactivated vaccines, with the development of SARS-Cov-2 mutants, mRNA especially bivalent mRNA vaccine showed more broadly and strongly protective function against Omicron^26^. Therefore, the combination of FTC, drug use, and enhanced vaccination would play an important role especially in certain period, for instance, the busy Chinese Spring Festival migration season.

## Conclusion

FTC can decrease the deaths as an anti-pandemic, precise prevention method targeting the older population during the initial outbreak of Omicron and could be a cost-effective strategy, along with enhanced immunity level and drug use. Therefore, more ICU beds, highly effective vaccines, and Paxlovid drug should be prepared as part of the FTC implementation.

## Data Availability

All data produced in the present study are available upon reasonable request to the authors

## References

1. Cosar B, et al. SARS-CoV-2 mutations and their viral variants. Cytokine Growth Factor Rev 63:10–22 (2022).

2. Araf Y, et al. Omicron variant of SARS-CoV-2: genomics, transmissibility, and responses to current COVID-19 vaccines. J Med Virol 94:1825–1832 (2022).

3. Daily briefing: “Out of control” Omicron threatens China. Nature Briefing, 28 March 2022.

4. Suzuki R, et al. Attenuated fusogenicity and pathogenicity of SARS-CoV-2 Omicron variant. Nature 603:700–705 (2022).

5. Zhang J, et al. Clinical characteristics of COVID-19 patients infected by the Omicron variant of SARS-CoV-2. Front Med 9:912367 (2022).

6. Linas BP, et al. Projecting COVID-19 mortality as states relax nonpharmacologic interventions. JAMA Health Forum 3:e220760 (2022).

7. Lin YF, et al. Impact of combination preventative interventions on hospitalization and death under the pandemic of SARS-CoV-2 Omicron variant in China. J Med Virol (2022).

8. Liu J, Liu M, Liang WN, Perspectives: the dynamic COVID-zero strategy in China. China CDC Weekly 4: 74–75(2022)

9. Wang Y, et al. Assessing the feasibility of sustaining SARS-CoV-2 local containment in China in the era of highly transmissible variants. BMC Med 20:442 (2022).

10. Cai J, et al. Modeling transmission of SARS-CoV-2 Omicron in China. Nat Med 28:1468–1475 (2022).

11. Kenyon C. Flattening-the-curve associated with reduced COVID-19 case fatality rates: an ecological analysis of 65 countries. J Infect 81: e98–e99 (2020).

12. Strålin K, et al. Mortality in hospitalized COVID-19 patients was associated with the COVID-19 admission rate during the first year of the pandemic in Sweden. Infect Dis 54:145–151 (2022).

13. Dai MY, et al. Patients with lung cancer have high susceptibility of COVID-19: a retrospective study in Wuhan, China. Cancer Control. 27:1073274820960467 (2020).

14. Liu M, et al. Lessons learned from early compassionate use of convalescent plasma on critically ill patients with COVID-19. Transfusion 60:2210–2216 (2020).

15. Leng Y, et al. Minimized glycemic fluctuation decreases the risk of severe illness and death in patients with COVID-19. J Med Virol 93:4060–4062 (2021).

16. Zhao L, et al. The clinical and bioinformatics analysis for the role of antihypertension drugs on mortality among patients with hypertension hospitalized with COVID-19. J Med Virol 94:4727–4734 (2022).

17. Fan Y, et al. SARS-CoV-2 Omicron variant: recent progress and future perspectives. Signal Transduct Target Ther 7:141 (2022).

18. Zhong J, et al. Heterologous booster with inhaled adenovirus vector COVID-19 vaccine generated more neutralizing antibodies against different SARS-CoV-2 variants. Emerg Microbes Infect 11:2689–2697 (2022).

19. China Health Statistics Yearbook. http://www.stats.gov.cn/tjsj/ndsj/2021/indexeh.htm. (2021).

20. http://www.nhc.gov.cn/yzygj/s7653p/202203/a354cb3151b74cfdbac6b2e909f311e6.shtml.

21. https://www.moh.gov.sg/covid-19/statistics.

22. Stuart ASV, et al. Immunogenicity, safety, and reactogenicity of heterologous COVID-19 primary vaccination incorporating mRNA, viral-vector, and protein-adjuvant vaccines in the UK (Com-COV2): a single-blind, randomised, phase 2, non-inferiority trial. Lancet 399:36–49 (2022).

23. Chalkias S, et al. A bivalent omicron-containing booster vaccine against COVID-19. N Engl J Med 387:1279–1291 (2022).

24. Cheng ZJ, et al. Public health measures and the control of COVID-19 in China. Clin Rev Allergy Immunol 18:1–16 (2021).

25. Hammond J, et al. Oral nirmatrelvir for high-risk, nonhospitalized adults with COVID-19. N Engl J Med 386:1397–1408 (2022).

26. Fang Z, et al. Bivalent mRNA vaccine booster induces robust antibody immunity against Omicron lineages BA.2, BA.2.12.1, BA.2.75 and BA.5. Cell Discov 8:108 (2022).

