## Supplemental Figure 1-3 for "Is it possible to flatten-the-curve after the initial outbreak of Covid-19? A data-driven modeling analysis for Omicron pandemic in China"

### Slide 1
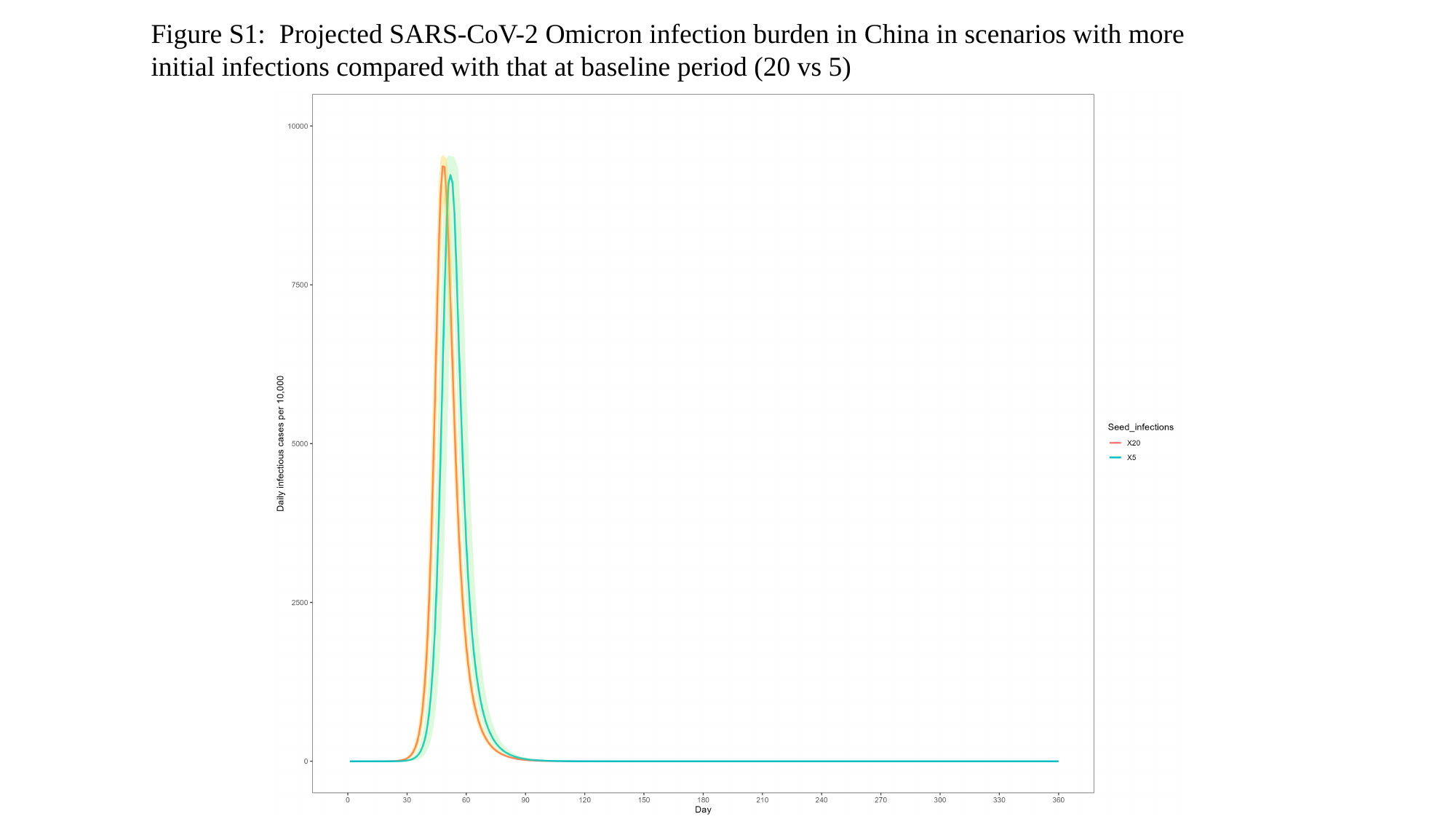

Figure S1: Projected SARS-CoV-2 Omicron infection burden in China in scenarios with more initial infections compared with that at baseline period (20 vs 5)

### Slide 2
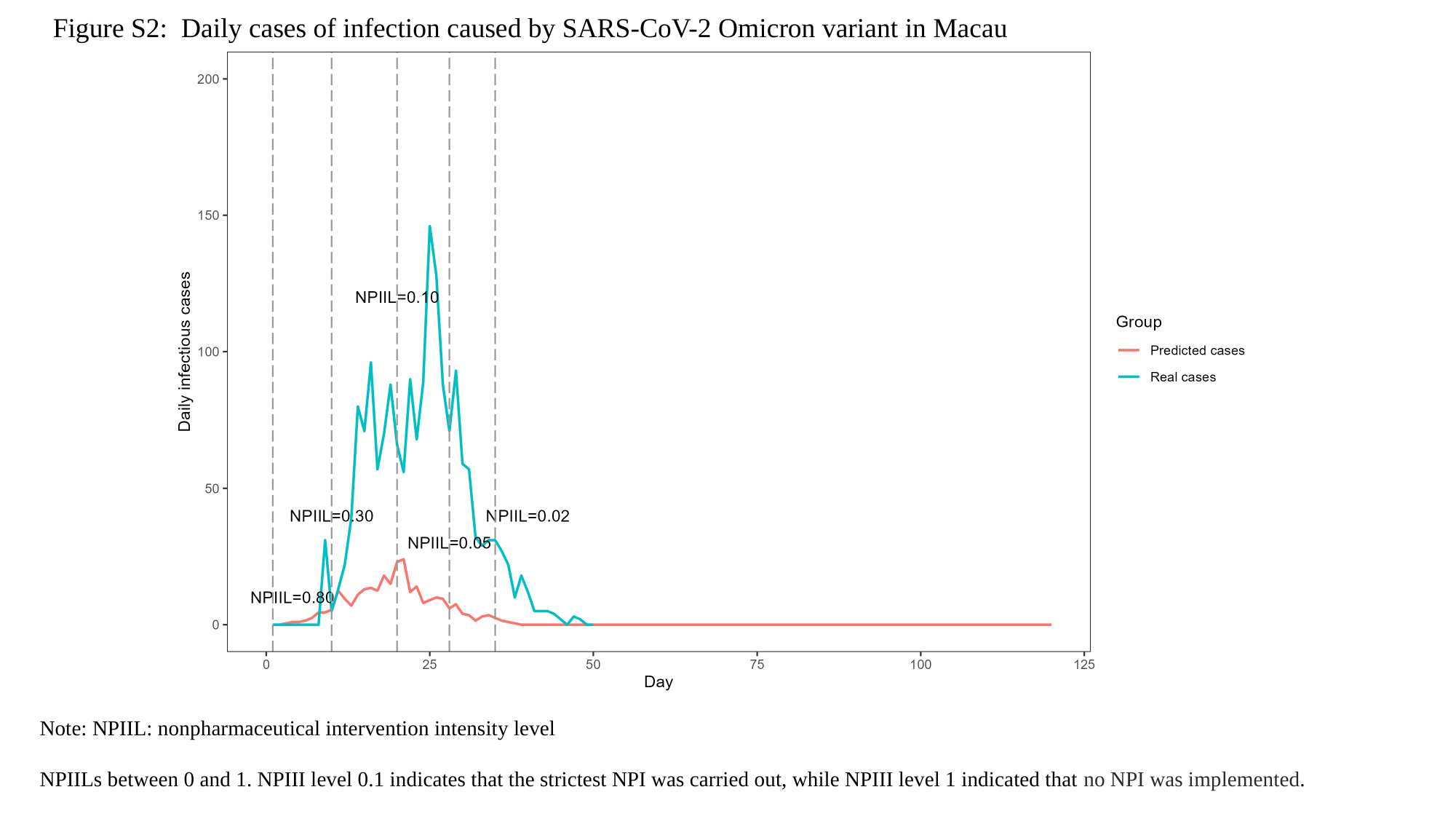

Figure S2: Daily cases of infection caused by SARS-CoV-2 Omicron variant in Macau
Note: NPIIL: nonpharmaceutical intervention intensity level
NPIILs between 0 and 1. NPIII level 0.1 indicates that the strictest NPI was carried out, while NPIII level 1 indicated that no NPI was implemented.

### Slide 3
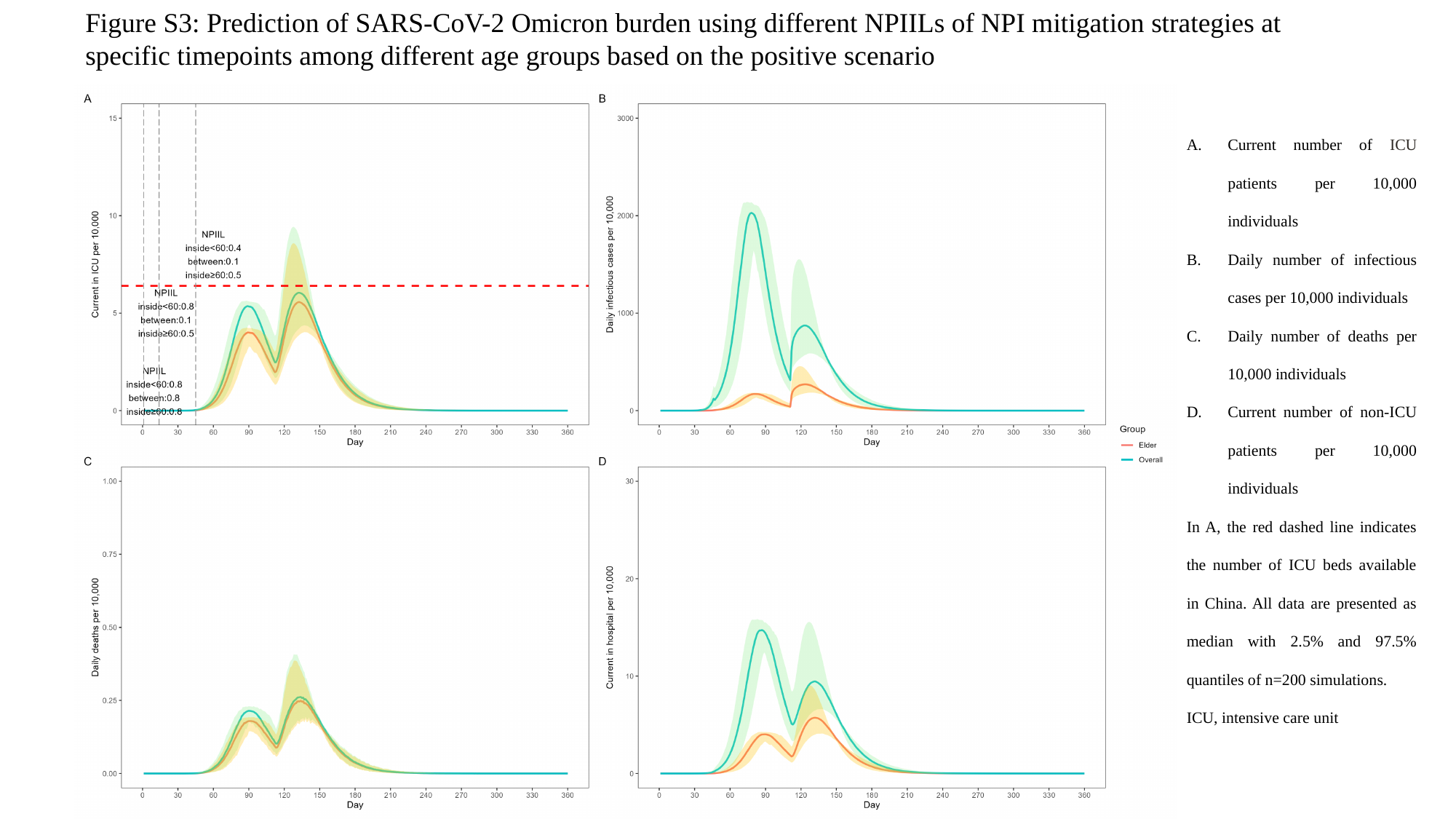

Figure S3: Prediction of SARS-CoV-2 Omicron burden using different NPIILs of NPI mitigation strategies at specific timepoints among different age groups based on the positive scenario
Current number of ICU patients per 10,000 individuals
Daily number of infectious cases per 10,000 individuals
Daily number of deaths per 10,000 individuals
Current number of non-ICU patients per 10,000 individuals
In A, the red dashed line indicates the number of ICU beds available in China. All data are presented as median with 2.5% and 97.5% quantiles of n=200 simulations.
ICU, intensive care unit
